# N95 respirators, disposable procedure masks and reusable cloth face coverings: total inward leakage and filtration efficiency of materials against aerosol

**DOI:** 10.1101/2020.11.24.20237446

**Authors:** Scott Duncan, Paul Bodurtha, Syed Naqvi

**Affiliations:** Defence Research and Development Canada- Suffield Research Centre, Chemical Biological Assessment and Protection Section, Medicine Hat, Alberta, Canada

## Abstract

Humans expel physiological particles continuously through normal respiratory activities such as breathing, talking, coughing and sneezing; a portion of these are aerosol in the size range <5.0 µm. Misconceptions exist on how to best implement face coverings as an effective preventive health measure against potentially infectious respiratory generated aerosol. The aim of this study was to characterise the performance of face coverings against aerosol when worn by individuals, and to quantify the maximum aerosol penetration through the material used in the construction of each mask. The former addresses their use as a means of possible protection against aerosol present in the environment and the latter having relevance to filtration and reducing human generated aerosol from reaching the environment. Face covering performance was assessed by measuring the total inward leakage of aerosol through the mask material and face seal. Aerosol penetration was measured on swatches of material taken from the face covering. An inert polydisperse charge-neutralized NaCl aerosol, with a distribution ranging from 0.023 µm to 5 μm in diameter, was used for the experiments.

Total inward leakage tests were completed to assess the protection factor for nine variations of face coverings, including seven reusable cloth masks, of which six were homemade and one was commercially manufactured, and two styles of disposable procedure masks, one with ear loops and one with ties. Our results have shown that face coverings in general provide the wearer only limited protection against aerosol in the environment. All reusable cloth face coverings obtained a practical protection level of less than 2. The performance of the disposable procedure masks varied from 1.7 to 3.6. The mean practical protection level for the nine face coverings was 1.95 with a standard deviation of 0.89. Comparatively, a N95 respirator achieved a protection factor of 166. We have further shown that aerosol readily penetrates through most materials used in face coverings. Aerosol swatch penetration tests were completed on six different fabrics commonly available for reusable homemade face coverings, four different material systems comprised of multiple material types, eight different disposable procedure masks and the filtering material from three different N95 respirators. Maximum aerosol penetration through the six common fabrics varied from 39% to 91%; for systems comprised of multiple types of materials 4% to 23%; for materials used in disposable procedure masks 16% to 80%; and for filtering materials used in N95 respirators 1.0% to 1.9%. We also highlight that with the exception of some of the reusable cloth materials, penetration of particulates at 5 µm diameter, representing the minimum filtration efficiency that could be achieved against droplets, was insignificant; the six common fabrics showed penetration from 1% to 44%; the fabric systems comprised of multiple types of materials <0.9%; the materials used in disposable procedure masks <0.9% to 6%; and the filtering materials used in three different N95 respirators <0.9%. The observations from this study directly demonstrate that face coverings may be optimized by incorporating high filtration efficiency materials in their construction. Face coverings with an enhanced level of filtration would be of benefit in circumstances where SARS-CoV-2 may be present in the aerosol of infected individuals to reduce aerosol emission from respiratory activities penetrating through into the environment.

## Introduction

On 11 March 2020 the World Health Organization (WHO) declared that a global pandemic was underway caused by the severe acute respiratory syndrome coronavirus 2 (SARS-CoV-2) (1). Most countries at the national, provincial/state and municipal/county level have implemented measures requiring the closure of non-essential businesses and where feasible, work-from-home policies or minimal staffing with controlled public access, in an effort to slow the rate of virus transmission through their populations. Some countries have enforced a complete lockdown, only permitting the public to access certain essential services (2). At this juncture nearing six months into the pandemic, most countries have opened many aspects of their economy, including business, recreation and education. The centre point of most doctrine provided by governments to safeguard health and wellness of individuals is social or physical distancing, some form of face covering usage, practicing good hand washing hygiene and disinfecting commonly touched surfaces. These measures are meant to address the three principal routes for pathogen transmission between humans: i) contact transfer of body fluid from an infected individual to the hands of another person and then self-contamination to areas with accessible mucosa; ii) droplet spray from an infected person coming in direct contact with areas of exposed mucosa of another individual, referred to as droplet transmission; and iii) fine particles produced through normal respiratory events such as breathing, talking, coughing and sneezing by an infected individual which stay suspended in the air for tens of minutes to hours that may contain a viable viral load and subsequently inhaled by another individual, referred to as airborne transmission (3). Evidence suggests that each of these routes plays a role in disease transmission but for any given pathogen the relative importance may vary (4-6).

New thoughts are being expressed on the nature of aerosolised disease transmission resulting from particles expelled during normal respiratory activities that question the longstanding advice recommending a requirement of only 1 meter separation between health care workers and infected persons (7, 8). The discussion is largely centred round the World Health Organisation’s relatively rigid definition for airborne versus droplet routes of transmission, which uses a particle diameter cut-off of less than 5 µm to demarcate airborne transmission and greater than 5 µm for droplet transmission (9). The WHO recommends contact and droplet precautions for SARS-CoV-2 predicated on maintaining 1 m separation from the infected individual (10, 11) whilst the Centers for Disease Control and Prevention recommends 2 m physical distance (12). Bahl *et al*. (7) concludes that there are limited studies to adequately inform on the appropriate physical distance and increasing evidence that following only precautions for droplets is not sufficient for the SARS-CoV-2 virus.

It is well understood from the literature that normal physiological respiratory activities such as sneezing, coughing, talking and even breathing, expel particles into the local environment where individuals interact (13-18). The range of particle sizes, the shape of the size distributions and particularly the particle number concentrations is somewhat disparate, but this is due more to the methodological approaches and technologies used to measure the particles, and to some extent, the variability of the subject populations used in the studies. Recent evidence suggests that the expulsion of particles in some respiratory activities is more appropriately described in the manner of a multiphase turbulent high momentum cloud, or puff, comprised of warm, moist air that envelops a wide range of droplet and aerosol sizes, delaying the rate of evaporation and extending the distance travelled (19-21). Many numerical studies suggest that distances aerosol and droplets may travel on ejection from the oral-nasal tract may reach 7-8 m (19, 20, 22-26). Moreover, numerous studies have established that aerosol expelled during normal respiratory activities can contain infectious influenza virus (3, 27-34). Recently, a large study investigating the presence of influenza virus in exhaled breath containing aerosol (>0.05 µm and ≤5 µm) detected infectious virus in 39% of samples and influenza virus RNA in 76% of samples collected during 30 minutes of normal tidal breathing (35). The stability of virus in the environment will play an important role in the disease transmission process. Temperature and relative humidity in particular can drastically affect the rate of decay of virus survival in aerosol (36). Other studies have demonstrated that influenza virus is detectable in the air and on surfaces in health care settings (37-40, 41). A laboratory study investigating the stability of SARS-CoV-2 in aerosol suspensions showed that it remained viable in aerosol 1-3 µm for up to 16 h (42). Findings from a similar study have confirmed a half-life of 177 min for SARS-CoV-2 in simulated saliva aerosol 1-3 µm in size at a relative humidity of 68-88% (43).

Many governments and health jurisdictions are promoting the wearing of face masks when out in public as a means to reduce the risk of exposure to SARS-CoV-2 under circumstances where individuals are involved in activities that require them to be in close proximity to one another. Recent work has postulated that airborne transmission is the dominant route for the transmission of SARS-CoV-2 (44). These authors conclude that the high transmission rates of SARS-CoV-2 in those countries that only implemented social distancing, quarantine and isolation as the primary preventive health measures demonstrates that, compared to countries where transmission rates were measurably lower and had also instituted an associated policy mandating the wearing of face masks in public, proves that SARS-CoV-2 is primarily transmitted as an airborne disease. Unfortunately, this study (44) suggests that facemasks block the passage of aerosol generated through normal respiratory activities from escaping to the environment and that they also prevent the inhalation of aerosols. Face masks are more likely to reduce the transmission of infectious disease (45). Other studies have made it clear that information is still lacking on whether SARS-CoV-2 is spread through aerosols (46).

Numerous studies have been completed, many recently, discussing filtration efficiency of certified respirators and improvised face coverings and their materials (47-55). Face coverings are not respirators. They have not been designed to any filtration or protection standards and the vast majority of individuals have not been trained in their proper use; there should be no implicit expectation of the level of protection that will be achieved when they are worn. The current construct round the usage of face coverings is to minimise the number of respiratory generated droplets expelled into the environment by those wearing the mask as well as protect wearers against large droplets produced by others (3, 56). The role that face coverings play in protecting wearers from aerosol (particles <5 µm in diameter) has been given only limited consideration (55). The protection performance of a face covering must be considered both in terms of the filtration efficiency of the filtering material used in its construction and the inward leakage associated with the fit of the mask to the face of the wearer. The concept that masks play a dual role, protecting the external environment from an individual and the individual from the external environment, has been discussed previously (45). Davies *et al*. (47) observed that the number of microorganisms expelled through a surgical face mask and homemade facemask following coughing was 15% and 21.5% respectively, of the total measured without a mask, showing that face mask materials reduce but may not prevent aerosol from penetrating through to the environment. A recent study on the aerosol filtration efficiency of common fabrics used in homemade cloth masks using a particle size distribution from 0.01 µm to 6 µm found that for a single layer of fabric the filtration efficiency varied from 5% to 80% for particles <0.3 µm in diameter and 5% to 95% for particles >0.3 µm in diameter (48). Filtration efficiencies for fabrics improved when multiple layers were used and when using a specific combination of different fabrics. The protection performance of face coverings such as disposable procedure masks that do not fit tightly to the face will be dominated by the total inward leakage due to gaps in the fit (48, 49) and indeed fit has been demonstrated to play a critical role in the ability of a mask or respirator to protect the wearer from exposure to particulates in the environment (54). The WHO (57) are currently advising a fabric mask composition of at least three layers to take into consideration the possible use of lesser quality and more variable cotton sources such as t-shirts and handkerchiefs, based on testing results cited in their guidance document (50, 51).

N95 filtering facepiece respirators (FFRs) are certified under National Institute for Occupational Safety and Health (NIOSH) 42 CFR 84 regulations (58), and as such the penetration of non-oil-based particulates through the filtering material, at 0.3 µm aerodynamic diameter, cannot exceed 5%. Balazy *et al*. (59) found that MS2 (bacteriophage) penetration through one N95 FFR was below 5% whilst for a second model it was at 5.6% between particle diameters 40-80 nm. Lee et al. (49) conducted a particle size selective (0.04-1.3 µm) total inward leakage study to assess the protection factor of four N95 FFRs and nine surgical procedure masks. They found that the geometric mean protection factor (PF) for the N95 respirators was 21.5 and only 2.4 for three representative surgical masks of those evaluated. The authors further noted that 29% of the N95 respirators and 100% of the surgical masks obtained PFs <10 (49). Other studies also have shown a failure of N95 respirators to achieve a PF greater than 10 in 14% to 26% of those tested (60, 61). It is evident that irrespective of the filtration efficiency of the filtering media, the level of protection that is obtained by a user wearing a mask or respirator is overwhelmingly dictated by the fit against the face, and any gaps if present, will significantly reduce the level of protection and concomitantly increase the likelihood of inhalational exposure.

In this study we investigate N95 FFRs, disposable procedure masks and reusable homemade cloth masks against aerosol in the size range 0.02 µm to 5 µm. This aerosol size range includes the most penetrating aerodynamic particle diameter for both conventional filtration materials and electrostatic filtration mediums. It is also appropriate for assessing that fraction of an aerosol distribution that may be associated with airborne transmission (<5 µm) according to WHO guidelines (9). Likewise, the penetration of particulates 5 µm in diameter represents the minimum filtration efficiency that could be achieved against droplets, with higher filtration efficiency being achieved for those of a larger diameter. From our work we have determined the practical protection level, based on total inward leakage, afforded to individuals when wearing these face coverings. We also completed aerosol swatch penetration tests on the material used in the construction of each face covering to quantify their filtration efficiency for aerosol in the same size range. We have shown that face coverings in general provide the wearer only limited protection against aerosol in the environment. We have further shown that aerosol (<5 µm) readily penetrates through most materials used in face coverings. With the exception of some of the reusable cloth materials, the penetration of particulates (droplets) >5 µm is insignificant. Thus, materials with a high filtration efficiency are considered critical to reduce the amount of aerosol that may penetrate through face covering materials and enter the environment.

## Materials and methods

### Face coverings and respirators

This study investigated fifteen face coverings and three models of N95 FFRs from different manufacturers. Of the fifteen face coverings, six were home-made and constructed from the following materials (the text in brackets denotes how they are identified in this study): two layers of cotton fabric (CC); two layers of silk fabric (SS); one layer each of quilt batting and cotton fabric (QC); three layered system comprised of one layer of furnace filter (3M™ Filtrete furnace filter, Merv 13) between an outer and inner layer of cotton (CEFC); four layered system comprised of two layers of quilt batting between an outer and inner layer of cotton (C2QC); bi-laminate in-house technical research material (ITRM). These face coverings were washable and thus considered reusable. The remaining nine face coverings were commercial-off-the-shelf (COTS) products, which included one reusable cloth mask and eight disposable procedure masks. The COTS reusable cloth mask was a three layer system constructed from cotton, a PM 2.5 filter insert and rayon (CFIR) (Weddingstar, Medicine Hat, Canada)). The PM 2.5 filter was a multi-laminate consisting of a two non-woven layers (exterior), two melt blown polymer layers with electrostatic properties for high filtration efficiency (interior) and one activated carbon layer (middle). Three additional fabric systems, also representative of types that could be used in homemade face coverings, were assessed for filtration efficiency only: two layers of polyester fabric (PP); two layers of nylon (NN); two layers of quilt batting (QQ). The eight disposable face coverings were of the type typically used in hospital/clinical settings and many commercial establishments and obtained from the following suppliers: Model HT Jiuzhuo Disposable Face Mask (HT-Jiuzhuo), Jiangsu Haitong Jiuzhuo Ecology Technology Co. Ltd., China; Model YL 5005 AlphaAir (YL5005-AA), AlphaProTech Inc., Salt Lake City, USA; Model PG4-1200 PrimaGard (PG4-1200), priMED Medical Products Inc., Edmonton, Canada; Model 836185 Disposable Face Mask (Henan Liwei), Henan Liwei Biological Pharmaceutical Co. Ltd., Jiaozuo City, China; Model PG4-2001 PrimaGard Surgical Mask Level 1 Barrier, Tie, (PG4-2001) priMED Medical Products Inc., Edmonton, Canada; Vanch Disposable Medical Face Mask (V-DMFM), 2020 Beifa Group Co. Ltd., Ningbo China; PG4-1273 PrimaGard Level 3 Barrier (PG4-1273), priMED Medical Products Inc., Edmonton, Canada; PG4-2331 PrimaGard Level 1 Barrier (PG4-2331), priMED Medical Products Inc., Edmonton, Canada.

The three models of FFR are NIOSH N95 certified and include the following: 3M™ model 9210 (fold style); Halyard Health, model FLUIDSHIELD 2 (duck bill style); North Safety Products model 7130N95 (cup style). The FFRs were chosen specifically for their differences in style and manufacturer, providing variability in respirator fit and filtration.

### Aerosol Swatch Penetration

The experimental aerosol swatch penetration set-up is shown in Figure 1. Located within the mixing chamber are two aerosol generators (TSI Model 8026, Shoreview, USA) and three fans positioned to maintain a steady uniform aerosol concentration of inert sodium chloride (NaCl) particles between 45,000-60,000 particles/cm^3^, permitting a reliable measurement of filtration efficiency of greater than 99.99%. A scanning mobility particle sizer (SMPS) spectrometer (TSI model 3080 with long differential mobility analyzer TSI model 3081 and ultrafine condensation particle counter TSI model 3776) was used to measure aerosol over the size range 0.023 - 0.67 µm, and an aerodynamic particle sizer (APS) spectrometer (TSI model 3321) was used to measure aerosol over the size range 0.5 - 20 µm. The aerosol distribution in the mixing chamber was polydisperse, extending over the range from ∼0.023 µm to ∼5 μm, with a geometric mean mobility particle diameter of 0.073 µm (geometric standard deviation of 1.69) corresponding to an aerodynamic particle diameter of 0.28 µm. The use of NaCl particles as a challenge aerosol, and its associated size distribution, has been shown to be an appropriate simulant to represent size ranges of both bacteria and viruses (47, 49). Balazy et al. (59) has shown similar penetration of MS2 virus and sodium chloride particles through N95 FFRs and surgical masks. Moreover, the use of NaCl aerosol is an accepted challenge medium for filtration testing of N95 FFRs by NIOSH (62) to quantify and qualify the performance of respirators.

**Figure 1:**
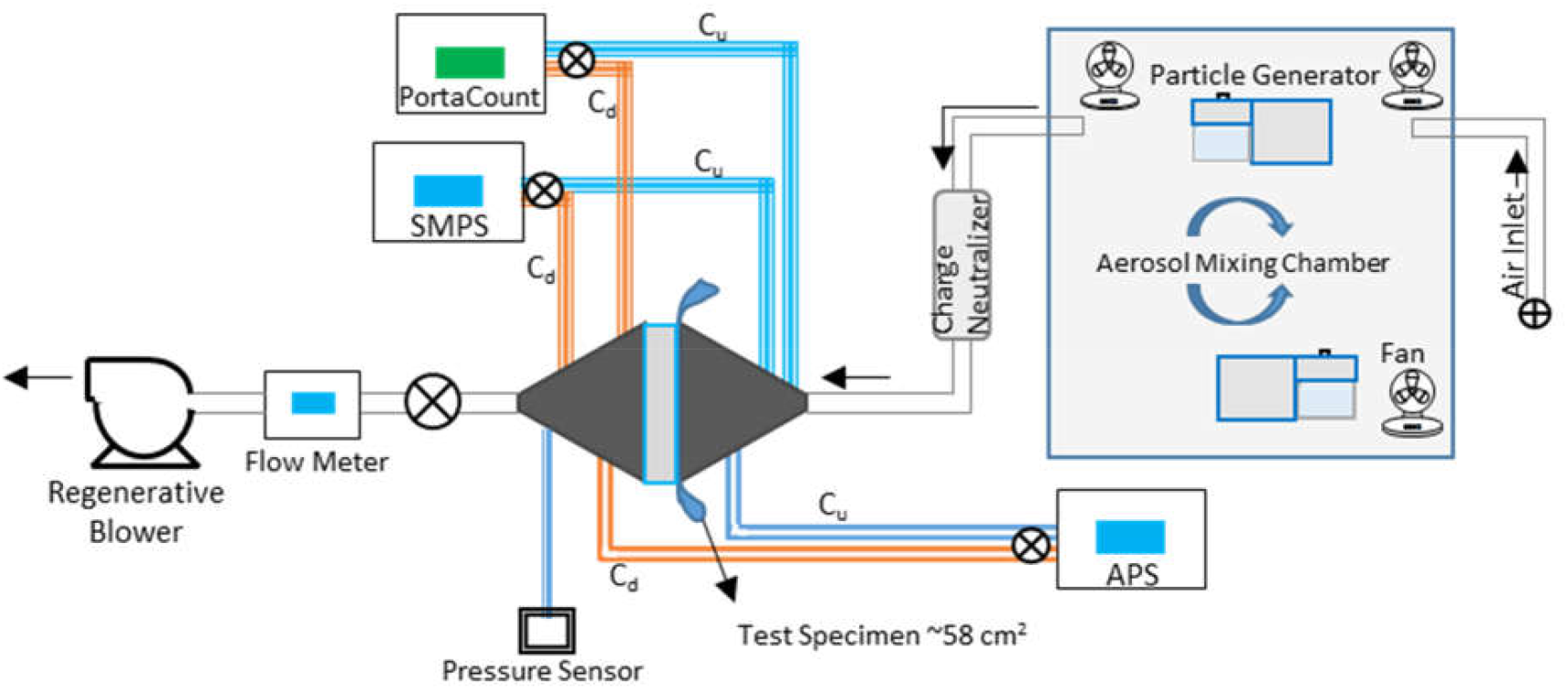
Schematic of aerosol swatch penetration set-up. A polydisperse NaCl aerosol is generated in the mixing chamber and pulled through the charge neutralizer and swatch test rig using a regenerative blower. Air flow is regulated using the flow meter. Aerosol concentration, upstream and downstream, is measured with the SMPS/APS and PortaCount as shown.

The aerosol size distribution, relative humidity (RH) and temperature were measured to be in compliance with US Code of Federal Regulations (63) for filtration testing of N95 FFRs (25 ± 5°C and 30 ± 10% RH), as referenced by NIOSH (62). Aerosol from the mixing chamber is drawn through a charge neutralizer (TSI Model 3054) in compliance with (63), neutralizing the surface charge on the particles to a Boltzmann equilibrium state, and then drawn through the fabric in the sample holder. A regenerative blower (GAST model R1102K-01), needle valve and digital flow meter (TSI Model 5330-2) were used to adjust the flow rate.

The fabric sample holder consisted of a central housing in two parts joined by a clamp to hold the material sample in place with no leaks. The holder was constructed in two sizes, with an inside diameter of 8.6 and 5.8 cm, allowing for an exposed sample area of 58.1 and 26.4 cm^2^, respectively. All face coverings were tested on the larger sample holder at a flow rate of 17 L/min simulating a respiratory rate for a light metabolic work level as defined by the International Organization for Standardization (ISO) (64). The associated face velocity through the face coverings at this air flow rate was 4.87 cm/s. The smaller sample holder was used to test the filter insert of the CFIR face covering, and was tested at a flow rate (7.8 L/min) that generated the same face velocity as the larger sample holder. The N95 FFRs also were tested on the larger sample holder at similar flow rates to the face coverings. Face velocities measured through the material were somewhat lower (1.73, 2.85 and 1.10 cm/s) due to their three dimensional form resulting in a larger surface area, and their more complex shape, which impacted how they were retained and sealed in the test rig. The differential pressure (ΔP) across the fabric sample materials was measured with an Ashcroft CXLdp Pressure transducer (Part# CX4MB21015IWL-XRH and CX4MB210P25IWL-XRH) to determine breathing resistance.

The aerosol penetration through the fabric sample was determined by measuring the aerosol concentration upstream and downstream of the fabric in the sample holder. This was accomplished through a series of valves that allowed the SMPS and APS to continually switch between upstream and downstream measurements. Number count versus aerosol size data collected with these instruments was analyzed using a commercial software package (TSI Model 390069 Data Merge Software Module) that converts the SMPS particle mobility size distribution to aerodynamic diameter units and merges this with the APS distribution to obtain a composite aerodynamic diameter distribution over the size range 0.023-5.0 µm. The resulting composite distribution is fitted to a mathematically rendered log-normal distribution curve. An example of the output from the Data Merge Software is illustrated in Figure 2 showing the measured SMPS and APS data, and the best-fit log-normal distribution curve for the composite distribution. Aerosol penetration through the face covering and N95 FFR materials was calculated by dividing the fitted upstream log-normal distribution curve by the downstream log-normal distribution curve. Where appropriate we have also calculated the extrapolated log-normal distribution curve beyond the range of our data to facilitate further understanding of aerosol penetration through the materials. In addition, as a second means of verification of aerosol penetration though the materials investigated, a condensation nucleus particle counting instrument (TSI PortaCount model 8020) was plumbed into the sample holder upstream and downstream of the material sample to measure aerosol over the size range from 0.02 to ∼1 µm. Custom software (Royal Military College of Canada, Kingston) was used to provide real-time aerosol penetration data.

**Figure 2:**
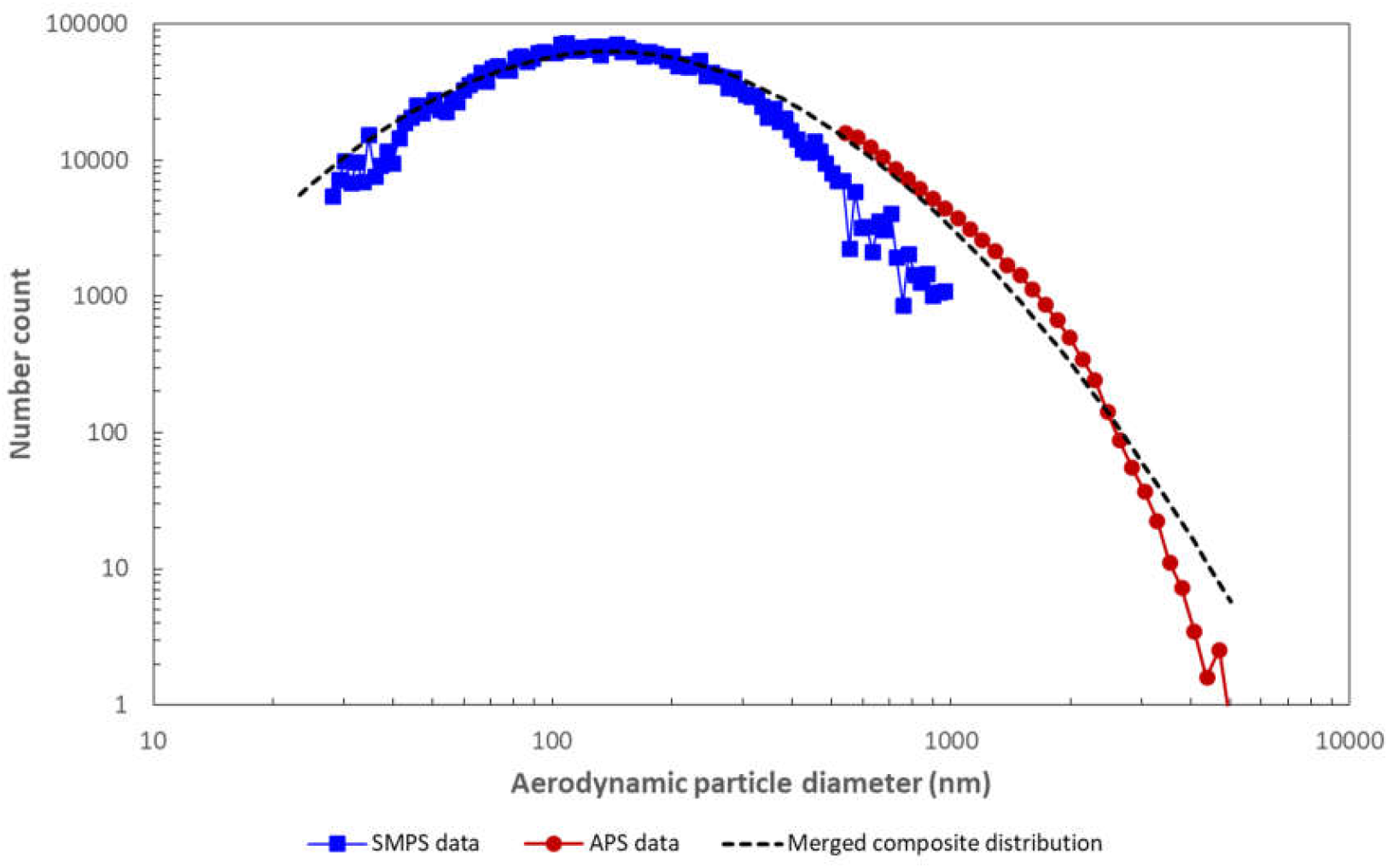
Example of the output from the Data Merge Software (TSI) showing the measured SMPS and APS data, and the best-fit log-normal distribution curve for the merged composite distribution.

### Face covering/FFR total inward leakage

This test measured the total inward leakage of aerosol through the face covering or N95 FFR when worn by a test subject and exposed to an aerosol. Here total inward leakage refers to penetration of aerosol though gaps where the face covering or respirator contacts the face as well as through the filtering material used in the construction of the face covering or FFR. The same aerosol particle size distribution and concentration was used for the total inward leakage testing as was used for the aerosol penetration swatch test. The total inward leakage test was conducted in accordance with Canadian Standards Association (CSA) Z94.4-18 (65) quantitative fit test (QNFT), and included the seven activities involving different head and face motions, each 30 s in duration. Aerosol measurements were obtained with the TSI PortaCount model 8038 inside of, and external to, the face covering/FFR. To measure the aerosol concentration inside the face covering/FFR a rivet-style sampling probe (TSI Model 8025-N95Adaptor Kit) was inserted through the material at a location between the mouth and nose of the subject and connected to the PortaCount via tubing, along with tubing for the external aerosol sampling.

The test matrix for the total inward leakage tests is provided in Table 1. A pool of eleven test subjects were recruited for this test from Defence Research and Development Canada (DRDC) Suffield Research Centre (Ralston, Canada) and were chosen based on obtaining a cross-section of both males and females, with a wide distribution of age, height and weight. Before testing the N95 FFR eight subjects were trained and successfully quantitatively fit tested using the TSI PortaCount model 8038, according to CAN/CSA Z94.4-18 (65). In order to be issued a specific N95 FFR, CSA Z94.4-18 and the US Occupational Safety and Health Administration (OSHA) (66) require that users of N95 FFRs achieve a minimum fit factor (FF) of at least 100. Note, for FFRs the QNFT FF test only measures particulate leakage at the face seal and excludes particulate penetration through the filter. Statistical analyses included one-way analysis of variance pair-wise comparisons (Tukey means comparison test) at P=0.05 significance level (Origin Pro v8 statistical package).

**Table 1:** Test matrix for the total inward leakage tests. HM refers to homemade reusable cloth masks, and COTS refers to commercial-off-the-shelf masks (manufactured masks such as CFIR, disposable procedure masks and N95 FFRs). Subjects wore each face covering/FFR once.

| Face covering/Filtering Facepiece Respirator | ID | Type | Number of subjects/tests |
| --- | --- | --- | --- |
| Cotton fabric – 2 layers | CC | HM | 8 |
| Silk – 2 layers | SS | HM | 5 |
| Quilt batting/Cotton – 2 layers | QC | HM | 5 |
| Cotton/Furnace Filter/Cotton – 3 layers | CEFC | HM | 5 |
| Cotton/2x quilt batting/cotton – 4 layers | C2QC | HM | 5 |
| Bi-laminate in-house technical research material | ITRM | HM | 5 |
| Cotton/PM2.5 filter insert/cotton – 3 layers | CFIR | COTS | 5 |
| Disposable procedure mask – ear loop | Henan Liwei | COTS | 8 |
| Disposable procedure mask - ties | PG4-2331 | COTS | 5 |
| N95 FFR | 3M 9210 | COTS | 8 |

## Results

Table 2 lists various fabrics and materials that were evaluated for aerosol penetration. Also provided is the aerosol penetration for three models of N95 FFR for reference. Fabric protection factors (FPF) are listed for both instrument measurement techniques employed in the study, the PortaCount and the SMPS/APS systems. FPF describes the reduction in aerosol concentration due to the filtering efficiency of the material. For the SMPS/APS data, FPFs were first calculated for each particle size bin from the measurement of the upstream number count divided by the corresponding downstream number count, and then an overall FPF was calculated as the harmonic mean of the FPFs from all the size bins. Similarly, the FPF for the PortaCount was calculated from measuring the upstream number count divided by the downstream number count. The difference in the FPF measurement between the two systems is likely due to the difference in the particle size distribution measured; the largest diameter particle measured by the PortaCount is ∼1 μm whilst the SMPS/APS system measures particles up to 20 μm in diameter. Our data only considered particles up to 5 µm in diameter as few were observed above this size. The maximum penetration and the most penetrating particle diameter (at maximum penetration) was calculated from the merged log-normal composite distribution curve-fit of the measured SMPS and APS data. The penetration at 5 µm was calculated directly from the upstream and downstream particle count data measured by the APS. To limit the counting statistical error on the penetration measurements at 5 µm the cumulative particle count was measured over a period of 180 seconds, for both the upstream and downstream. The limit of detection (LOD) was calculated to be <0.009.

**Table 2:** Aerosol penetration data for various fabrics and materials that were evaluated in this study. Also provided is the aerosol penetration for three N95 FFR materials. The difference in the penetration and practical protection levels measured by the PortaCount and the SMPS/APS instruments reflects the fact that the PortaCount only measures particles between 0.01-1 µm whilst the value for SMPS/APS data is calculated over the particle diameter range 0.023-5.0 µm.

| Fabric System | Instrument | Fabric Protection Factor (FPF) | Maximum penetration (most penetrating particle diameter $\mu\text{m}$)** | Penetration at 5 $\mu\text{m}$*** | Face Velocity (cm/s) | Pressure Drop (Pa) |
| --- | --- | --- | --- | --- | --- | --- |
| *Polyester fabric (2 layers) (PP) | PortaCount | 1.5 | - | - | 4.87 | 24.5 |
|  | SMPS/APS | 1.58 | 0.91 (0.53) | 0.32 |  |  |
| *Quilt batting (2 layers) (QQ) | PortaCount | 3.20 | - | - | 4.87 | 22.6 |
|  | SMPS/APS | 4.57 | 0.39 (0.38) | 0.04 |  |  |
| *Nylon fabric (2 layers) (NN) | PortaCount | 2.00 | - | - | 4.87 | 375.7 |
|  | SMPS/APS | 3.18 | 0.87 (0.92) | 0.01 |  |  |
| Bi-laminate in-house technical research material (ITRM) | PortaCount | 131.1 | - | - | 4.87 | 260.8 |
|  | SMPS/APS | 96.6 | 0.02 (0.17) | < 0.009 |  |  |
| Cotton fabric (2 layers) (CC) | PortaCount | 1.40 | - | - | 4.87 | 39.2 |
|  | SMPS/APS | 1.90 | 0.82 (0.33) | 0.44 |  |  |
| Cotton/Furnace Filter/Cotton (3 layers) (CEFC) | PortaCount | 5.10 | - | - | 4.87 | 49.1 |
|  | SMPS/APS | 14.2 | 0.25 (0.06) | < 0.009 |  |  |
| Silk fabric (2 layers) (SS) | PortaCount | 2 | - | - | 4.87 | 109.8 |
|  | SMPS/APS | 2.5 | 0.68 (0.53) | 0.03 |  |  |
| Quilt batting/Cotton (2 layers) (QC) | PortaCount | 2.1 | - | - | 4.87 | 38.2 |
|  | SMPS/APS | 2.1 | 0.71 (0.60) | 0.09 |  |  |
| Cotton/2x quilt batting/cotton (4 layers) (C2QC) | PortaCount | 8.7 | - | - | 4.87 | 48.0 |
|  | SMPS/APS | 11.2 | 0.22 (0.20) | < 0.009 |  |  |
| Cotton/PM 2.5 filter insert/rayon (3 layers) (CFIR) | PortaCount | 66.1 | - | - | 4.87 | 109.8 |
|  | SMPS/APS | 73.04 | 0.048 (0.08) | < 0.009 |  |  |
| PM 2.5 filter insert only (CFIR) | PortaCount | 60.9 | - | - | 4.87 | 55.9 |
|  | SMPS/APS | 69.1 | 0.058 (0.070) | < 0.009 |  |  |
| Disposable procedure mask | PortaCount | 8.43 | - | - | 4.87 | 50.0 |
| **(Henan Liwei)** | SMPS/APS | 17.66 | 0.18 (0.09) | < 0.009 |  |  |
| **Disposable procedure mask (YL5005-AA)** | PortaCount | 10.1 | - | - | 4.87 | 54.9 |
|  | SMPS/APS | 14.5 | 0.19 (0.08) | 0.02 |  |  |
| **Disposable procedure mask (PG4-1200)** | PortaCount | 7.4 | - | - | 4.87 | 39.2 |
|  | SMPS/APS | 19.35 | 0.16 (0.07) | < 0.009 |  |  |
| **Disposable procedure mask (HT Jiuzhuo)** | PortaCount | 1.93 | - | - | 4.87 | 21.6 |
|  | SMPS/APS | 2.3 | 0.80 (0.13) | 0.06 |  |  |
| **Disposable procedure mask (PG4-1273)** | PortaCount | 9.5 | - | - | 4.87 | 24.5 |
|  | SMPS/APS | 15.4 | 0.21 (0.07) | < 0.009 |  |  |
| **Disposable procedure mask (Vanch)** | PortaCount | 8.9 | - | - | 4.87 | 42.7 |
|  | SMPS/APS | 13.3 | 0.23 (0.09) | < 0.009 |  |  |
| **Disposable procedure mask (PG4-2001)** | PortaCount | 11.7 | - | - | 4.87 | 26.7 |
|  | SMPS/APS | 15.4 | 0.17 (0.06) | < 0.009 |  |  |
| **Disposable procedure mask (PG4-2331)** | PortaCount | 9.6 | - | - | 4.87 | 21.6 |
|  | SMPS/APS | 14.5 | 0.21 (0.07) | < 0.009 |  |  |
| **N95 North Safety 7130 FFR** | PortaCount | 230 | - | - | 2.84 | 52.5 |
|  | SMPS/APS | 206 | 0.019 (0.074) | < 0.009 |  |  |
| **N95 Halyard FS2 FFR** | PortaCount | 252 | - | - | 1.10 | 45.1 |
|  | SMPS/APS | 707 | 0.010 (0.074) | < 0.009 |  |  |
| **N95 3M 9210 FFR** | PortaCount | 230 | - | - | 1.73 | 54.9 |
|  | SMPS/APS | 243 | 0.016 (0.043) | < 0.009 |  |  |

\*Fabric systems only evaluated for aerosol filtration efficiency, not as face coverings.
\*\* The maximum penetration (and most penetrating particle diameter) was calculated from the merged log-normal composite distribution curve-fit of the measured SMPS and APS data.
\*\*\* The penetration at 5 µm was calculated directly from the upstream and downstream particle count data measured by the APS. To limit the counting statistical error on the penetration measurements at 5 µm the cumulative particle count was measured over a period of 180 seconds, for both the upstream and downstream. The limit of detection (LOD) was calculated to be < 0.009.

Each curve in Figures 3 to 6 showing the aerosol penetration as a function of aerodynamic particle diameter is the log-normal composite distribution obtained from a best-fit to the merged SMPS and APS data, as discussed above and illustrated in Figure 2.

**Figure 3:**
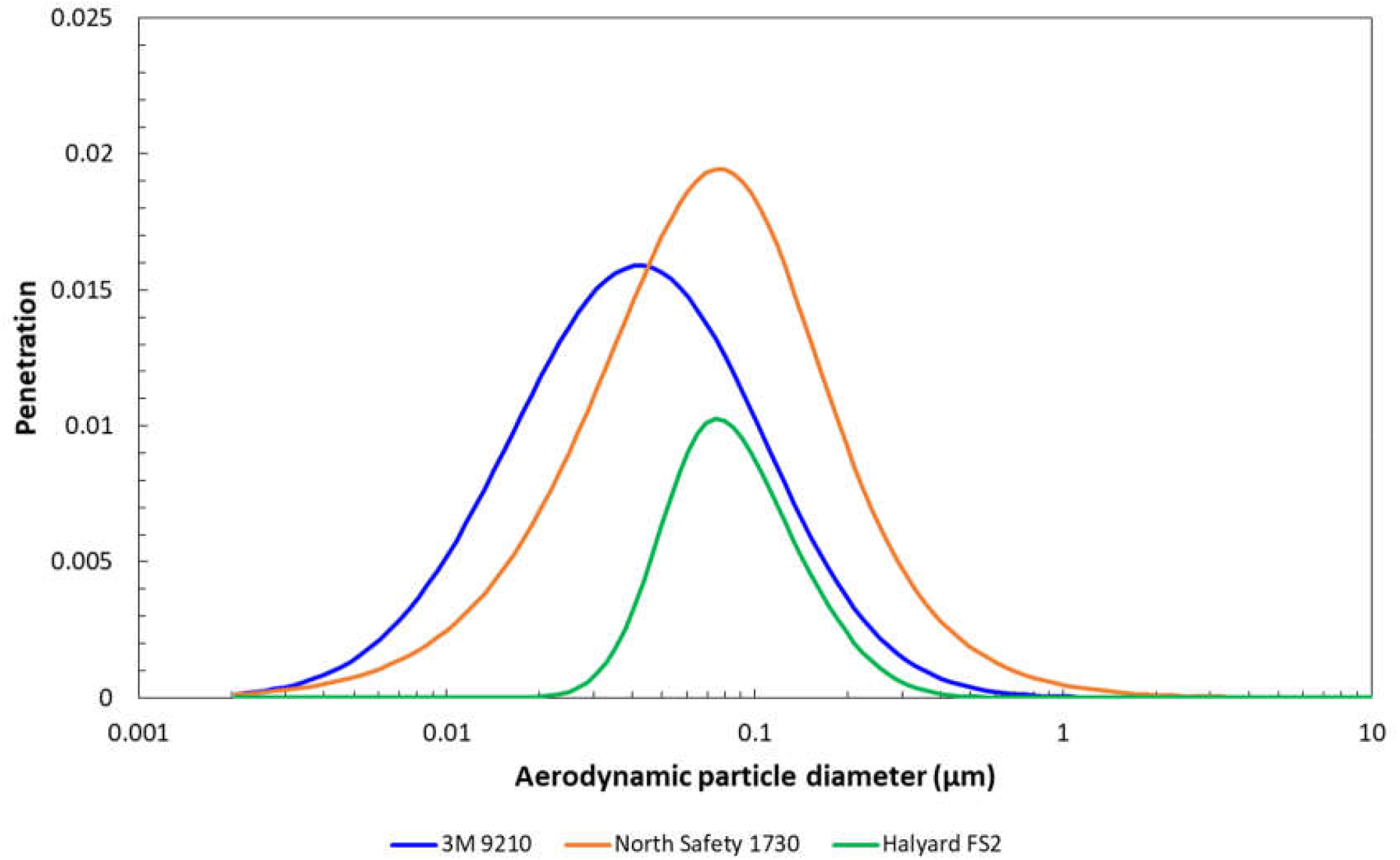
Penetration profile as a function of aerodynamic particle diameter for the filter material of three N95 FFRs. The curves shown are the merged log-normal composite distribution curve-fit obtained from the SMPS and APS data. Aerosol penetration was collected at a nominal flow rate of 17 L/min. Note the y-axis scale maximum is 0.025. A penetration value of 0.025 equates to 2.5% of aerosol penetrating through the material.

**Figure 4:**
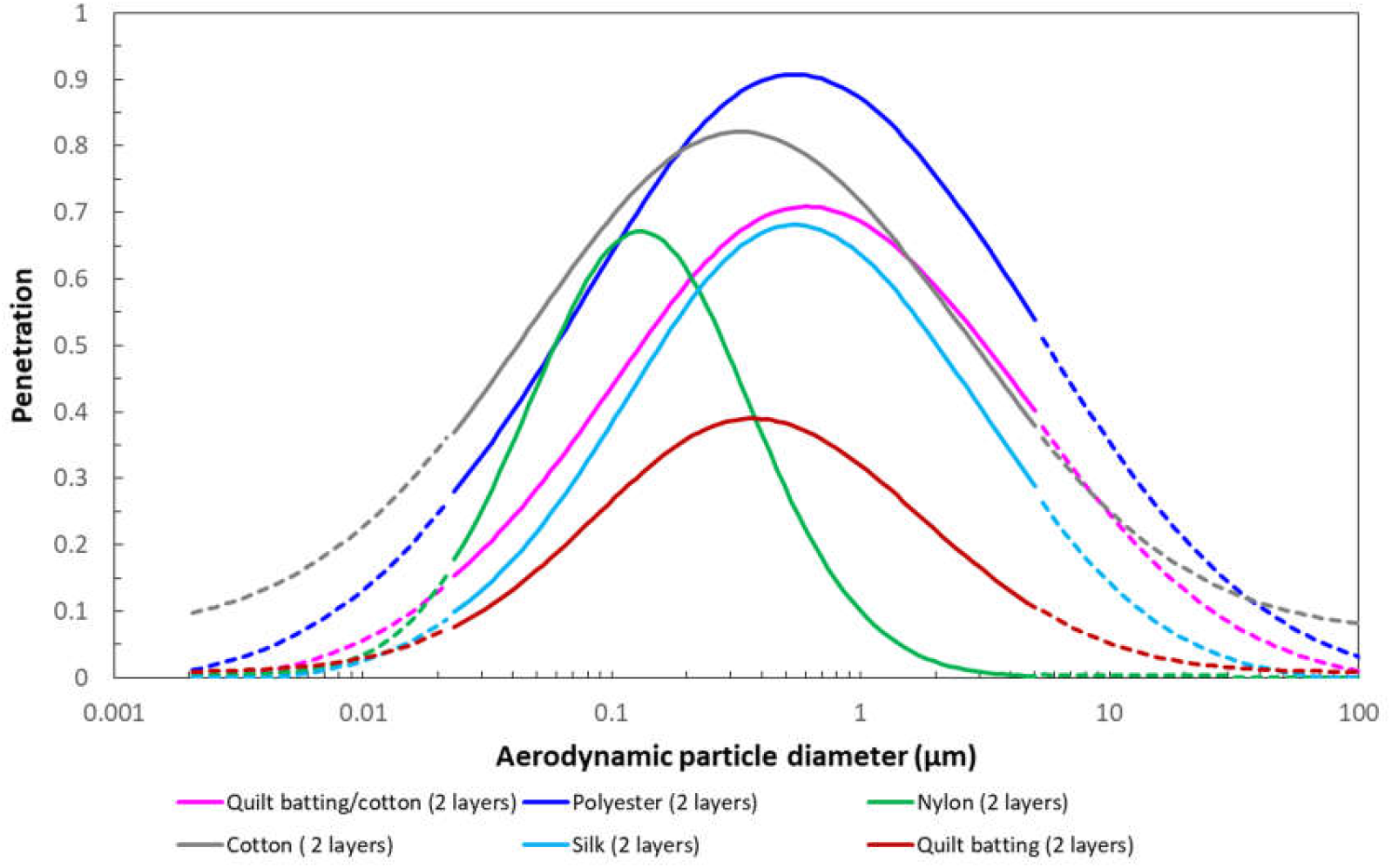
Penetration profile as a function of aerodynamic particle diameter for common fabrics used in reusable cloth face coverings. The solid segment of the curves are the merged log-normal composite distribution curve-fit obtained from the actual measured SMPS and APS data. Dashed lines show extrapolated penetration values for the particle size distribution beyond the experimental measurement range of 0.023 to 5.0 µm investigated. A penetration value of 1 equates to 100% of aerosol penetrating through the material.

**Figure 5:**
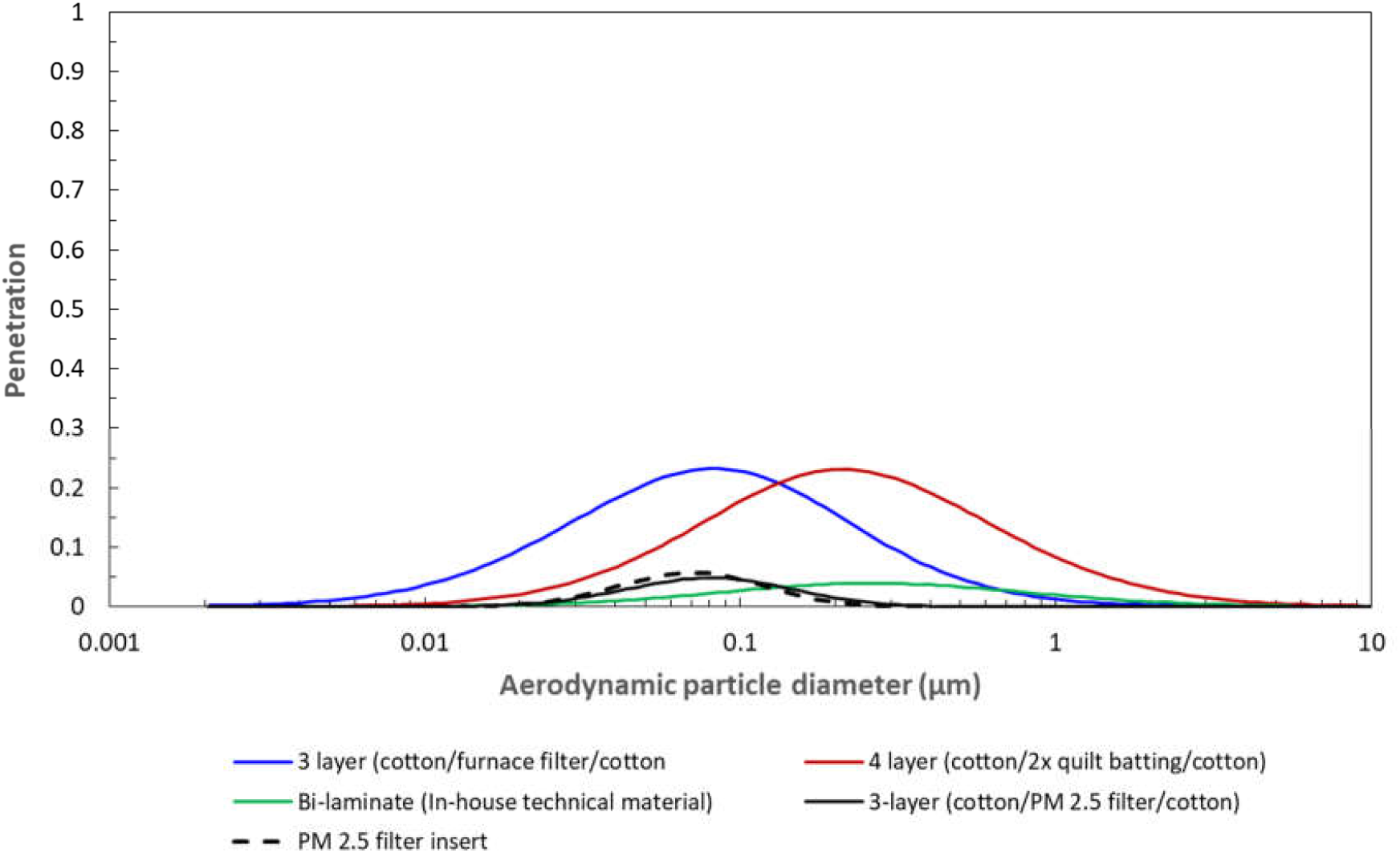
Penetration profile as a function of aerodynamic particle diameter for four multi-layer material systems that may be used in reusable cloth face coverings. Solid lines are the merged log-normal composite distribution curve-fit obtained from the actual measured SMPS and APS data. A penetration value of 1 equates to 100% of aerosol penetrating through the material. The aerosol penetration through only the PM 2.5 filter insert is shown separately as a dashed line.

**Figure 6:**
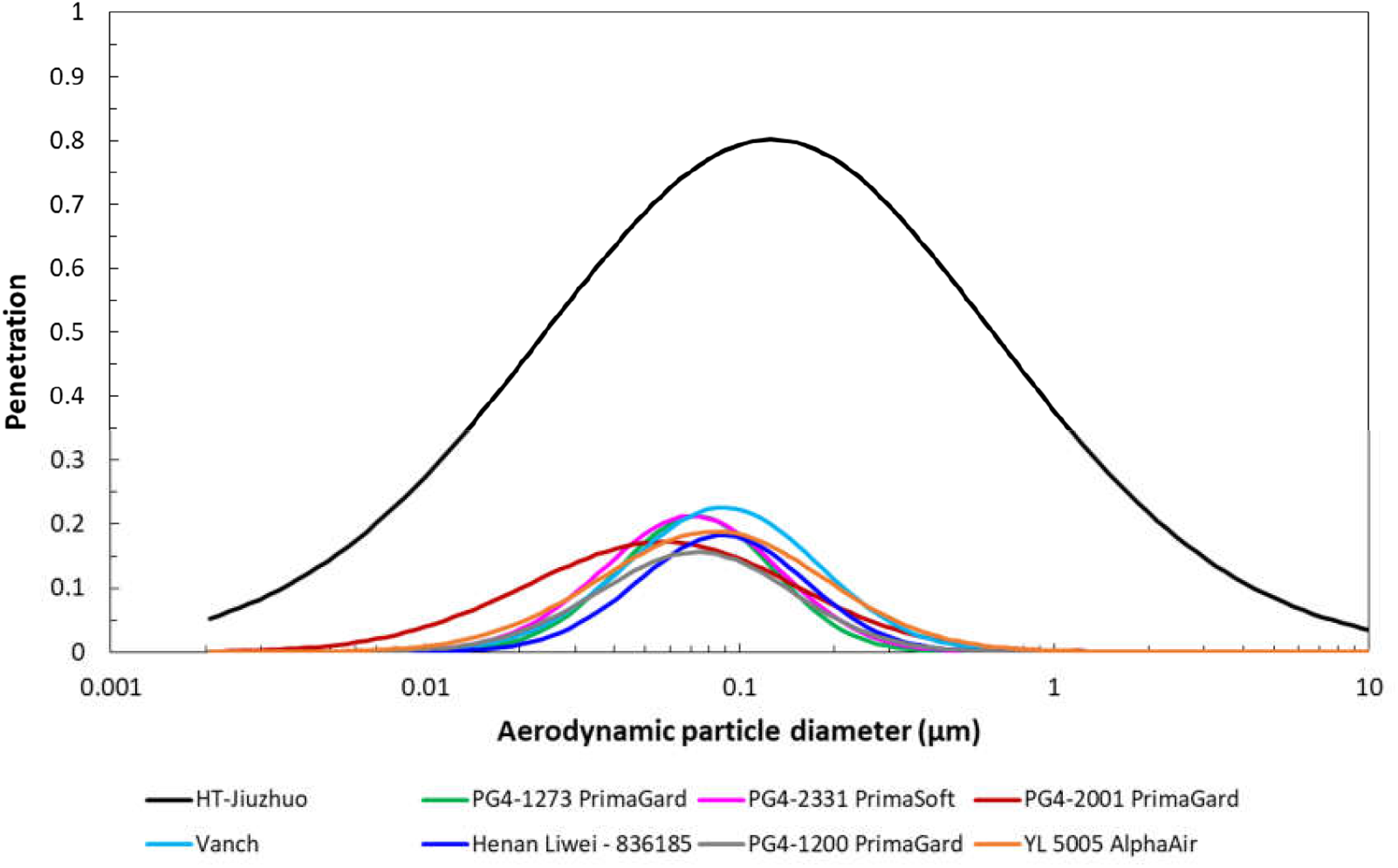
Penetration profile as a function of aerodynamic particle diameter for eight disposable procedure mask material systems. Solid lines are the merged log-normal composite distribution curve-fit obtained from the actual measured SMPS and APS data. A penetration value of 1 equates to 100% of aerosol penetrating through the material.

### Aerosol penetration

#### N95 FFR materials

Aerosol penetration as a function of aerodynamic particle diameter for three N95 FFRs is shown in Figure 3. The maximum penetration through the N95 filtering material varied from 0.010 to 0.019. This was expected given the low flow rate used here to simulate breathing at a light work rate. The most penetrating aerodynamic particle diameter at maximum penetration occurred over the range 0.043 to 0.074 µm. This is similar to what others have found (59, 67, 68, 69). Penetration at 5 µm was shown to be at the limit of detection (<0.009) for all three N95 FFRs. Pressure drop across the N95 FFR materials was similar, varying from 45.1 to 54.9 Pascals. Each FFR was also tested for aerosol penetration and breathing resistance at a nominal flow rate of 85 L/min as required by NIOSH for N95 FFRs (63); total penetration was less than 5% at an aerosol diameter of 0.3 µm, and inhalation and exhalation breathing resistance was less than 35 mm H2O (343 Pa) and 25 mm H2O (245 Pa) respectively, confirming that the N95 FFRs met performance standards.

#### Face covering materials – common materials (2 layers)

Many different fabrics are being considered for reusable face coverings. Common materials that individuals may have available include cotton, nylon, polyester, silk, and quilt batting. Figure 4 presents the aerosol penetration as a function of aerodynamic particle diameter for five fabrics, each consisting of two layers, and a sixth combination material comprised of two different fabric layers. The solid segment of the curve represents the best-fit to the merged log-normal distribution obtained from the SMPS and APS experimental data over the range 0.023 to 5.0 µm. The dashed lines show the extrapolated distribution beyond this range based on a mathematical best-fit curve to the experimentally measured data; fits for the extrapolated data were obtained using curve fitting software (Origin Pro v8) at R^2^>0.99. Peak aerosol penetration values were found to vary from 0.91 for two layers of polyester to 0.39 for two layers of quilt batting. The most penetrating particle diameter varied from 0.13 µm through two layers of nylon to 0.60 µm through the two layer fabric combination of quilt batting and cotton. Penetration at 5 µm particle diameter varied from 0.01 through two layers of nylon to 0.44 through two layers of cotton. Pressure drop across the fabrics depended on the tightness of the weave and ranged from 22.6 Pa for two layers of quilt batting to 393.6 Pa for two layers of nylon.

#### Face covering materials – multi-layer systems

The aerosol penetration through four multi-layer material systems was investigated. These included a 3-layer system comprised of cotton/electret furnace filter/cotton (CEFC), 4-layer system consisting of cotton/2 layers quilt batting/cotton (C2QC), a bi-laminate in-house technical research material (ITRM) and a commercially available cotton/PM 2.5 filter insert/rayon (CFIR). The penetration characteristics of these material systems are provided in Table 2 and the aerosol penetration profile as a function of aerodynamic diameter is shown in Figure 5. The maximum aerosol penetration varied from a minimum of 0.04 for the ITRM with a most penetrating particle size of 0.25 µm to a maximum of 0.23 for both the 3-layer CEFC fabric system and the 4-layer C2QC fabric system, although the most penetrating particle size for the former was lower, 0.08 µm compared to 0.21 µm. The low aerosol penetration through the CFIR (0.048) is due specifically to the PM 2.5 filter insert between the outer and inner layers. For comparison, a separate aerosol penetration test was performed on only the PM 2.5 filter insert and is shown as a dashed line in Figure 5; the maximum penetration was 0.058. Aerosol penetration through the material system used in the construction of the COTS available CFIR mask was the lowest of the reusable cloth materials tested and similar to the ITRM (Figure 5) and N95 FFR materials (Figure 3). The small diameter of the most penetrating particle for the CEFC and CFIR materials is a reflection of their electrostatic properties. Penetration at 5 µm was shown to be at the limit of detection (<0.009) for all four multi-layer material systems. Pressure drop across the five fabric systems varied from 48 Pa for C2QC to 260.8 Pa for ITRM. The high pressure drop across the ITRM makes it very difficult to breathe through and this has a negative impact on the overall performance of a face covering made from this material.

#### Face covering materials – disposable procedure mask materials

There are many manufacturers of disposable procedure masks and most manufacturers offer numerous models. In this study we investigated disposable procedure masks distributed/manufactured by five different companies as well as three additional models from one of the companies, for a total of eight disposable procedure masks. Notably, as shown in Figure 6 aerosol penetration through seven of the eight mask materials was very similar, characterised by maximum aerosol penetration from 0.16 to 0.23 with an associated maximum penetrating particle diameter in the range 0.06 to 0.09 µm. Fabric protection factors varied from 13.3 to 19.4, with a mean of 15.6 and standard deviation of 1.14. Penetration at 5 µm was shown to range from 0.02 to < 0.009. One disposable procedure mask (HT Jiuzhuo) had dramatically higher maximum aerosol penetration of 0.80 with a slightly larger maximum penetrating particle diameter of 0.125 µm (Figure 6). In this case, the FPF was 2.3. Penetration at 5 µm was lower at 0.06 but still much higher than the other masks of this type.

### Respirator and face covering protection performance

Figure 7 shows the practical protection level (PPL) determined from total inward leakage tests on nine variations of face coverings, including seven reusable cloth masks and two styles of disposable procedure masks, one with ear loops (Henan Liwei) and one with ties (PG4-2331). These are compared to the protection factor of one of the N95 FFRs (3M 9210) listed in Table 2. All reusable cloth face coverings except the four-layer C2QC obtained a practical protection level of less than two. The performance of the disposable procedure masks showed some variation, with the ear loop style (Henan Liwei), which was a typical non-sterile issue mask, providing a PPL of 1.7. The disposable procedure mask with ties (PG4-2331) was a Level 1 barrier surgical mask and provided a PPL of 3.6. A one-way analysis of variance pair-wise comparison was completed on the nine face coverings. The result indicated that the disposable procedure mask with ties and the reusable C2QC cloth mask were similar in performance but each statistically significantly different from the other seven face coverings at P=0.05 significance level (Tukey means comparison test). The mean PPL for the nine face coverings was 1.95 with a standard deviation of 0.89. Notably, the face covering that incorporated a PM 2.5 filter into its design (CFIR) only provided a PPL of 1.8. This contrasts markedly with the filtration performance of the PM 2.5 filter by itself, which provided a FPF of ∼69. The principal reason for this discrepancy is that, like all face coverings investigated here, this version also did not seal to the face, leaving gaps through which aerosol could penetrate. Further, the PM 2.5 filter insert only covered the area in front of the mouth and nose, not the entire mask, thus the outer area of the mask consisted only of two layers of fabric. The C2QC face covering outperformed the other reusable cloth masks and was purpose designed by our research team with several improvements, including two layers of quilt batting, which had been shown to provide better filtration than other conventional materials (see Figure 4), a quilt batting face seal, integrated nose strip and adjustable elastic cording for a tighter fit. The protection factor for the N95 3M 9210 FFR, which had the smallest most penetrating particle size of the three FFRs investigated (0.043 µm) and a maximum penetration of ∼0.016 (1.6%) (see Figure 3), is shown in Figure 7 for comparison. At a value of 166, this is 45 to 140 fold higher than the face coverings.

**Figure 7:**
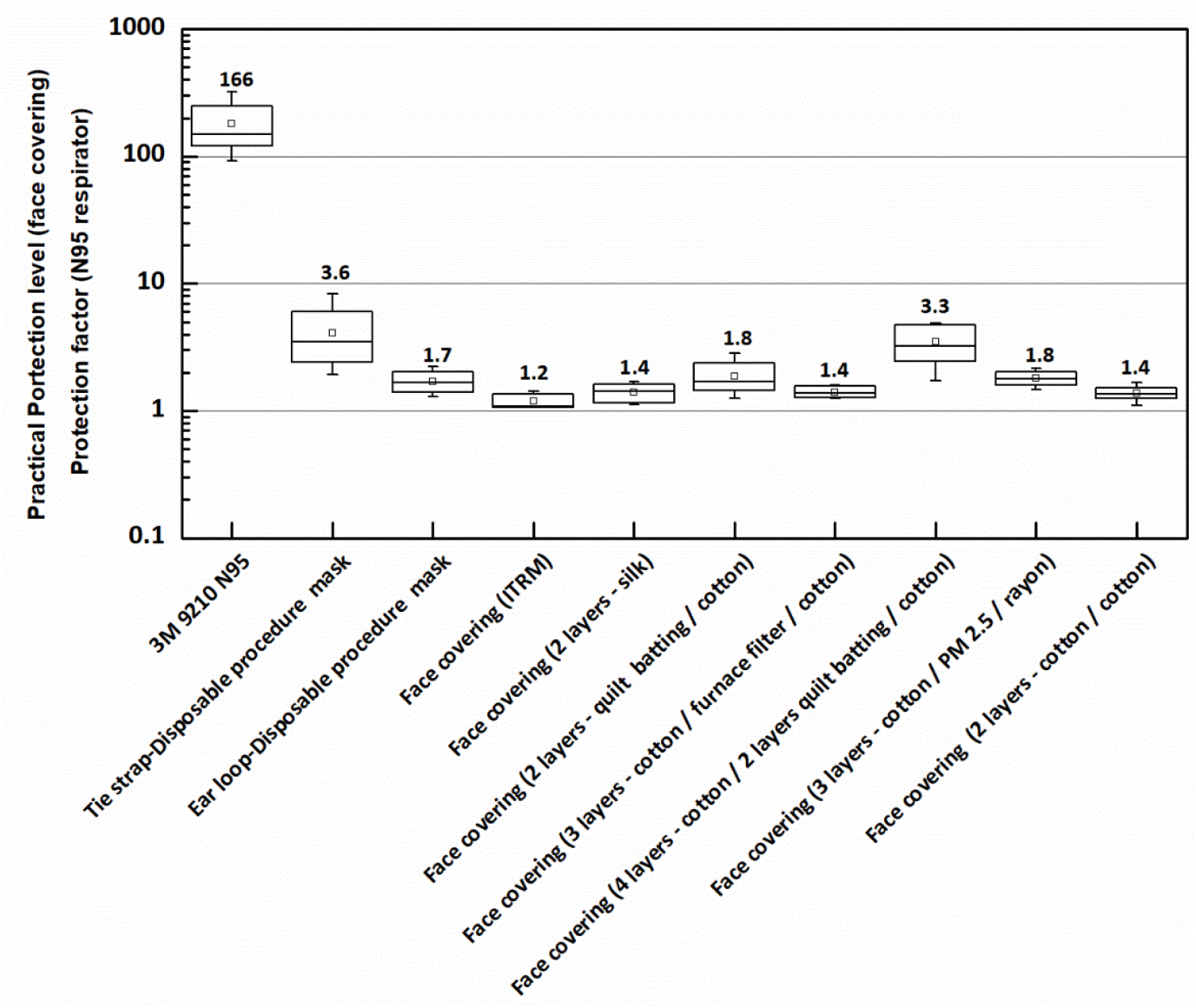
Practical protection level obtained for the various reusable cloth face coverings compared to the practical protection level for two different styles of disposable procedure mask and the protection factor obtained for a N95 FFR. The number shown above the marker denotes the geometric mean, also indicated by the small square marker; the line inside the box is the median, the upper and lower limit of the box are the 25^th^ and 75^th^ percentiles, and; the whiskers are the minimum and maximum values measured. The sample size for the N95 FFR, the disposable procedure mask with ear loops (Henan Liwei) and the reusable 2 layer cotton homemade face covering (CC) was n=8. For all other face coverings the sample size was n=5.

## Discussion

At this juncture the extent that aerosol and airborne transmission contributes to the spread of SAR-CoV-2 is not fully understood (70). Many studies however, have shown that infectious virus can be spread by aerosol (3, 27-35). Our study has clearly demonstrated the expected performance of face coverings such as commercially available disposable procedure masks and homemade reusable cloth masks as a preventive health measure (PHM) for reducing exposure to aerosols in the size range 0.023 to 5.0 µm. In contrast to elastomeric respirators, and even N95 FFRs, disposable procedure masks and reusable cloth masks do not provide a seal to the face. Aerosol present in the environment of the size range investigated here can readily pass through even small gaps of millimeter size, which may not be immediately evident to the eyes, but nevertheless represent openings on the order of one thousand times larger than the aerosol. Larger gaps are unhindered channels for aerosol to penetrate inside the face covering and potentially be inhaled. As expected, in terms of mask performance for total inward leakage, the N95 FFR significantly outperformed the face coverings with a mean PF of 166 (Figure 7). This corresponds to an overall FFR efficiency of 0.994 and may reduce an individual’s exposure to aerosol to <1% of the amount present in the environment. Substantially higher protective performance is achievable with the N95 FFR because, in this instance, i) each wearer was quantitatively fitted according to CAN/CSA Z94.4-18 (65) to obtain a PF ≥100 prior to the total inward leakage test being conducted, ii) the design of a FFR provides for a more effective face seal, and iii) the electret filtering material has a much higher filtration efficiency. A recent study by Hill *et al*. (54) showed that qualitatively fitting a KN95 and N95 FFR to a headform resulted in a FFR efficiency of only ∼0.40, but when the KN95 FFR was sealed to the headform with adhesive, the efficiency increased to 0.967.

The disposable procedure mask with ties (PG4-2331) and our customised face covering C2QC provided the highest PPL of all face coverings investigated at 3.6 and 3.3 respectively, which in turn corresponds to a mask efficiency of 0.72-0.70 and potential aerosol exposure of ∼30%. All remaining face coverings were found to have a PPL based on total inward leakage that was less than 2 (Figure 7). At this level, individuals would be exposed to more than 50% of aerosol in the environment. The mean PPL for all face coverings investigated, including the disposable procedure masks and reusable cloth masks, was only 1.95 with a standard deviation of 0.89. Lee *et al*. (49) found that the overall geometric mean protection factor of three models of surgical procedure masks selected at random from a larger group of nine models, was 2.4, which was nine times lower than the protection factor determined for N95 FFRs. A recent study by Sickbert-Bennett *et al*. (71) showed similar PPL results where they evaluated multiple brands of surgical and procedure face masks on one test subject, obtaining mean values of 3.5 and 1.6 respectively. Hill *et al*.’s (54) investigation also demonstrated that homemade face coverings provided as-worn filtration efficiencies of 15-40% (PPLs of 1.18 to 1.67). Given that disposable procedure masks and reusable cloth masks are, by simplest definition, a piece of material held in position over the nose and mouth, it may be stated reasonably confidently that a PPL of 2.0-2.5 is likely the upper level of performance that can be anticipated from basic designs of this nature. Noting that aerosol penetration through the material used in PG4-2331 and C2QC is 0.213 and 0.231 respectively (see Figure 5 and 6), which is significantly higher than that observed through the N95 FFR materials (see Figure 3), the improvement in the PPL relative to the other face coverings, albeit small, may be attributed to less leakage at the face. Accordingly, it is the lack of an efficient face seal that dominates the protection performance of non-certified face coverings.

Fabrics commonly available for homemade masks were characterised by high aerosol penetration (poor filtration efficiency) although they may still provide some level of filtration; peak aerosol penetration was shown to be in the 0.39 to 0.91 range (see Figure 4). Jung *et al*. (50) observed aerosol penetration through nonwoven and cotton materials of 0.45-0.77 and handkerchief material between 0.96-0.99. Others (51) found that common fabric materials and cloth masks were characterized by penetration levels ranging between 0.40-0.90 for aerosol 0.02 to 1.0 µm in diameter. Penetration at 5 µm was shown to vary considerably depending on the type of fabric material, ranging from as low as 0.01 through two layers of nylon to as high as 0.44 through two layers of cotton. There are ways to enhance the filtration of fabrics by combining them with other materials with better filtration efficiency (48), or using materials that are known to filter more effectively (e.g. furnace filter electrets and non-wovens (quilt batting)). Our materials CEFC and C2QC (Figure 5) showed a 40% improvement in filtration, reducing the maximum aerosol penetration to ∼0.23, compared to two stand-alone layers of quilt batting (QQ). Penetration at 5 µm was also reduced from 0.04 to less than <0.009, providing a marked improvement in droplet protection. Notably, the maximum penetration through the CFIR material was only 0.048. This is due to the PM 2.5 filter insert material incorporated into this reusable COTS mask, which as a stand-alone filter material, has a peak aerosol penetration of 0.058 (Figure 5), nearly on par with the filtration efficiency requirements of N95 FFRs. Surprisingly, the laminate materials used in the construction of most disposable procedure masks were quite effective at filtering aerosol in the size range 0.023 to 5.0 µm. Aerosol penetration was limited to 0.16 to 0.23 with the exception of one mask material where aerosol penetration was 0.80 (see Figure 6). Jung *et al*. (50) observed somewhat higher aerosol penetration through medical surgical and dental procedure mask materials of ∼0.58 and ∼0.30, respectively. It is particularly interesting to note that our bi-laminate in-house technical research material (ITRM), which was designed for non-respiratory applications, has a very high filtration efficiency (PF=131) on par with N95 FFRs, but a pressure drop ∼5 times higher than a N95 electret filter medium (261 Pa vs 55 Pa). It was included in this study to illustrate that using a material with a high filtration efficiency (Figure 5) but which also has a high pressure drop virtually guarantees poor mask performance. Such a high pressure drop severely restricts the passage of air through the material and essentially all of the air required for inhalation, as well as that expelled during exhalation, enters and exits through gaps where the mask meets the face. The PPL of the mask made from the ITRM was one of the lowest measured at 1.3 (Figure 7). Thus, high filtration efficiency properties for materials used in face coverings does not necessarily imply that a mask constructed from said material will provide a similarly high level of protection. This is further illustrated by the COTS reusable CFIR cloth mask with the PM 2.5 filter insert; it afforded no more than a PPL of 1.8 (Figure 7), despite the CFIR fabric itself achieving a peak aerosol penetration <0.05 (filtration efficiency of >95%). In this instance, the pressure drop across the CFIR material was less than half that of the ITRM material at 109.8 Pa, highlighting that even with significantly lower pressure drops the face seal remains the focal point of mask leakage.

This study focused on aerosol in the range 0.023 to 5.0 µm in diameter, which are produced during normal respiratory activities. It highlights a significant need to understand the application of PHMs such as face coverings in limiting the amount of aerosol that enters the human environment and reduce the risk of spreading a highly contagious virus like SARS-CoV-2 by airborne means. Our findings have shown that aerosol will penetrate through most common fabrics and materials used in face coverings to varying degrees and that the aerosol filtration efficiency of the materials used in the face coverings have little impact on the protective performance of the mask itself. Moreover, the face coverings themselves provide only a minimal level of protection to the wearer. Notwithstanding this, face coverings of the type discussed in this study, disposable procedure masks and reusable cloth masks, when worn properly to cover both the mouth and nose at all times, are a means to lessen the risk of wearers from exposure to aerosols generated by other individuals, and more importantly, to reduce the amount of aerosol produced by wearers via normal respiratory activities that may be expelled into the environment. Asadi *et al*. (55) have recently demonstrated both surgical masks and KN95 respirators reduce the outward particle emission rates by 90% and 74% during speaking and coughing, respectively, compared to when a mask is not worn. There is a benefit to using face coverings that incorporate materials with electrostatic filtration properties to enhance the overall filtration efficiency; the most penetrating aerodynamic particle diameter through electrostatic materials is <0.3 µm and often in the range 0.04-0.09 µm (59, 67, 68, 69). A number of materials in this study were observed to have a most penetrating aerodynamic particle diameter <0.09 µm, benefiting from the electrostatic effect; all of the N95 FFRs (0.043-0.074 µm, Figure 3), CEFC and CFIR (∼0.08 µm, Figure 5) and seven of the eight disposable procedure mask materials (0.06-0.09 µm, Figure 6). However, only the CFIR with its PM 2.5 filter insert had aerosol penetration comparable to N95 FFRs at <0.05. Noting that a single SARS-CoV-2 virion varies from 0.10-0.125 µm in size (42), reducing the overall aerosol penetration through mask materials used in PHMs whilst also limiting the penetration of aerosol to those as small as possible, will inherently translate to a smaller fraction of virion entrained in the aerosol and a corresponding reduction in the overall virion emission into the environment. The CFIR mask material was the only non-certified material to accomplish this effectively. Some studies consider only protection against virion-sized particulates (54). We point out however, volumetrically a 1 µm sized aerosol may potentially contain as many as 380 virion of 0.125 µm diameter (assuming face centred cubic packing). Thus, the filtering material in face coverings, and particularly whether it has electrostatic properties, becomes critical to manage the aerosol load produced during gatherings of people to address potential airborne disease transmission should some individuals in these groups be infected. It also follows that there is an obvious synergistic effect where if everyone in group settings were to wear a face covering made from a high filtration efficiency material the aerosol concentration that individuals would be exposed to would be proportionally that much lower, and the limitations of the protective performance of face coverings less of an issue. Accordingly, effective attenuation of respiratory generated aerosol is paramount and the biggest gain collectively to reduce the level of aerosol exposure comes from the filtration efficiency of the material used in their construction.

Haphazard implementation of PHM protocols, including wearing face coverings, has not been effective in eliminating the transmission of SARS-CoV-2. Unfortunately, collective acceptance and adherence to wearing face masks in public is not widespread in many jurisdictions. At the time of writing there have been numerous protests by certain factions of society demanding that governments withdraw mask wearing entirely. We certainly advocate mandatory wearing of face masks collectively in all public spaces to standardize the response to protective posture and address increased concerns round the potential airborne transmission of SARS-CoV-2. A recent study (72) found viable SARS-CoV-2 virus in the air up to 4.8 m from an infected individual. The genome sequence from the virus collected by air samplers was identical to that isolated from a nasopharyngeal swab taken from the infected individual. The authors of the study conclude that normal respiratory activities of infected persons may be a source of airborne transmission of the virus. As one would expect, N95 FFRs, given their certification requirements, were shown to provide the highest respiratory protection factor and the highest level of filtration efficiency. Thus, they are the best option to protect individuals from an aerosol environment in high risk settings, and the environment from individuals possibly shedding infectious virus in their respiratory aerosol. However, N95 FFRs are relatively expensive and supply chain shortages have made them difficult to source outside health care settings. The effectiveness of homemade face coverings as a PHM has been discussed in the literature (47, 73). Davies *et al*. (47) do not recommend the use of homemade face masks as a method of reducing transmission of infection from aerosols. Hill *et al*. (54) suggest caution when entering areas of high exposure risk wearing homemade face coverings and recommend using masks known to seal to the face. There is certainly merit in these statements due to the potential for a false sense of respiratory safety that may arise from mask use within the public.

However, the alternative of no one wearing a mask is considerably less desirable; some level of respiratory protection is better than none even with material penetration of 0.39-0.91 and PPLs that may be less than two. Worby *et al*. (73) provides a similar stance stating that although face masks have a limited protective effect, they can reduce total infections and deaths, and can delay the peak time of the epidemic. Our results have shown that aerosol in the size range 0.023 to 5.0 µm penetrates readily through most reusable cloth face covering materials, thus they may afford only minimal attenuation of aerosol produced from respiratory activities. It is this aerosol fraction that is most problematic for airborne transmission (42, 43, 72). Some reusable cloth face coverings also showed significant penetration at 5 µm (as high as 0.44), indicating potentially lower than desirable protection even against droplet transmission. Moreover, for some of the common fabrics shown in Figure 4 for example the systems with two layers quilt batting/cotton, two layers cotton and two layers polyester, particle penetration may extend well beyond an aerodynamic particle diameter of 50 µm as suggested by the extrapolated penetration profile. Although the aerosol penetration through most materials used in disposable procedure masks was shown to be <0.25 (providing >75% filtration efficiency), making them generally more effective at filtering in-mask respiratory aerosol than reusable cloth masks, some models are extreme outliers and are characterized by significantly higher penetration, as we observed in this study. This was an issue observed by Balazy *et al*. (59) as well. It is worth noting that most of the disposable procedure masks provided very effective filtration of 5 µm particulates (0.02 to 0.009) with the outlier mask reducing the penetration to 0.06. Unfortunately, it is not possible to know the filtration efficiency afforded by disposable procedure masks prior to donning them. Perhaps it is time that disposable procedure masks be certified to provide a required level of filtration efficiency more in line with, for example, PM 2.5 filters or N95 FFR materials. Accordingly, we suggest that the benefit of incorporating PM 2.5 filters, or other materials with similar filtration efficiency, into reusable homemade cloth masks to lessen the risk of aerosol particle emission through mask materials into the environment be made widely known by public health authorities. We further suggest that only wearing face coverings will not be sufficient to reduce the aerosol load in the air that will inevitably occur when groups of people gather in living, education and work environments where engineering controls may be insufficient to ensure effective air circulation and air exchange, or where groups of individuals are interacting at close quarters either indoors or outdoors (<1.0 m distance between individuals engaged in conversation, for example). Such situations represent potential high risk exposure conditions. Hence, in conjunction with mandatory wearing of face coverings, implementing the additional PHM requiring compulsory 2 m physical distance between individuals in all public spaces, will serve to reduce the human generated aerosol concentration in the environment even further through spatial dilution, and provide the greatest impact on reducing potential airborne mediated transmission of disease. Of course, good respiratory etiquette, hand hygiene, contact tracing, isolation of infected cases and immunization programmes must be in place.

## Conclusion

Face coverings such as disposable procedure masks and reusable cloth masks produced commercially or homemade can play an important role as a preventive health measure; some level of protection is more desirable than the alternative. In this study, which focussed on aerosol <5 µm, all reusable cloth face coverings obtained a practical protection level of less than 2. The performance of the disposable procedure masks varied from 1.7 to 3.6. The mean practical protection level for the nine face coverings was 1.95 with a standard deviation of 0.89. Comparatively, a N95 respirator achieved a protection factor of greater than 150. Maximum aerosol penetration through six fabrics commonly available for reusable homemade face coverings varied from 39% to 91%; for fabric systems comprised of multiple types of materials 4% to 23%; for materials used in disposable procedure masks 16% to 80%; and for filtering materials used in three different N95 respirators 1.0% to 1.9%. We also highlight that with the exception of some of the reusable cloth materials, penetration of particulates at 5 µm diameter, representing the minimum filtration efficiency that could be achieved against droplets, was insignificant; the six common fabrics showed penetration from 1% to 44%; the fabric systems comprised of multiple types of materials <0.9%; the materials used in disposable procedure masks <0.9% to 6%; and the filtering materials used in three different N95 respirators <0.9%. Our findings directly demonstrate that face coverings may be optimized by incorporating high filtration efficiency materials in their construction. Such face coverings that provide an enhanced level of filtration would be of benefit in circumstances where SARS-CoV-2 may be present in the aerosol of infected individuals to reduce penetration through the mask into the environment. Lessening the risk of exposure to potentially infectious aerosol would improve if there is collective implementation of more effective face coverings combined with physical distancing and other accepted preventive health measures.

## Data Availability

All relevant research data is provided in the paper as graphs and/or in table format.

## Acknowledgements

The authors thank Dr. Y. Lacroix MD, Dr. C. Willis PhD and Dr. A. Vallerand PhD for reviewing the manuscript and providing very helpful suggestions. The authors also would like to thank J4 Med Mat, CF Health Services Group and the National Research Council of Canada, METRO-M36, for providing N95 respirators, Liz Duncan for making the homemade cloth masks, the Royal Canadian Mounted Police for providing a TSI PortaCount 8038, Cheryl Jensen and Amber Stark for technical support to the study, the test subjects from DRDC Suffield Research Centre for their dedication and time, and Canadian Forces Base Suffield G4 Stores for the tent structure employed as the aerosol mixing chamber. This study was funded by Defence Research and Development Canada.

